# Real-time, interactive website for US-county level Covid-19 event risk assessment

**DOI:** 10.1101/2020.08.24.20181271

**Authors:** Aroon Chande, Seolha Lee, Mallory Harris, Troy Hilley, Clio Andris, Joshua S. Weitz

**Author notes:** Correspondence: Clio Andris & Joshua S. Weitz.

## Abstract

Large events and gatherings, particularly those taking place indoors, have been linked to multi-transmission events that have accelerated the pandemic spread of severe acute respiratory syndrome coronavirus 2 (SARS-CoV-2). To provide real-time, geo-localized risk information, we developed an interactive online dashboard that estimates the risk that at least one individual with SARS-CoV-2 is present in gatherings of different sizes in the United States. The website combines documented case reports at the county level with ascertainment bias information obtained via population-wide serological surveys to estimate real time circulating, per-capita infection rates. These rates are updated daily as a means to visualize the risk associated with gatherings, including county maps and state-level plots. The website provides data-driven information to help individuals and policy-makers make prudent decisions (e.g., increasing mask wearing compliance and avoiding larger gatherings) that could help control the spread of SARS-CoV-2, particularly in hard-hit regions.

## Main text

As of August 20, 2020, there have been over 5.6 million documented cases of SARS-CoV-2 in the United States, leading to more than 150,000 documented fatalities (World Health Organization, 2020). Despite large-scale efforts to suppress disease spread via lockdown orders and other non-pharmaceutical interventions including mask-wearing (Ferguson et al., 2020), there has been a resurgence of SARS-CoV-2 cases in the United States in late summer 2020, particularly in the South and West (New York Times, 2020b; The Atlantic Monthly Group, 2020). The rise of cases threatens public health, economic recovery, and the re-opening of K-12 schools as well as colleges/universities (Gottlieb & Strain, 2020; Levinson, Cevik, & Lipsitch, 2020).

The basic reproduction number and large fraction of asymptomatic cases represent challenges for controlling SARS-CoV-2 (Fauci, Lane, & Redfield, 2020). Early estimates of the basic reproduction number of SARS-CoV-2 range from 2.1 to 4.5 (Park et al., 2020), with current best estimates from the Centers of Disease Control (CDC) suggesting 2.5 (Centers for Disease Control and Prevention, 2020b). Early studies found that approximately half of cases may be via presymptomatic, mild, or asymptomatic transmission mild/asymptomatic (Lavezzo et al., 2020; Nishiura, Linton, & Akhmetzhanov, 2020). The absence of commonly associated symptoms (fever, cough, shortness of breath) may be more pronounced in younger individuals. In addition, effective isolation of symptomatic cases may increase the fraction of circulating cases that are mild/asymptomatic.

The strong and often undocumented spread of SARS-CoV-2 is exacerbated by large transmission incidents (Li et al., 2020), referred to as ‘super-spreading’ events (Kain et al., 2020). Super-spreading of SARS-CoV-2 has been documented in multiple, indoor events or large gatherings in which a single infector is putatively associated with the infection of dozens (or more) of individuals (Frieden & Lee, 2020; Liu, Eggo, & Kucharski, 2020; Xu et al., 2020). Large gatherings pose particular challenges for the prevention and spread of SARS-CoV-2. First, the number of potential interactions increases with gathering size (potentially as the square of the number of individuals in small groups where all individuals might be in contact). Second, follow-up contact tracing is problematic given the potential unknown nature of identifying close interactions. Third, the risk that one (or more) individuals is infected increases rapidly with group size; increasing the inherent risk of a potential exposure as groups increase in number. Although the first two challenges can be hard to quantify due to logistical and privacy reasons, this third category of risk is quantifiable, presents a gateway to action-taking, and should be communicated to the public at large.

In March 2020, one of us (JSW) developed a scenario-driven approach to assess the risk that one (or more) individuals in a group was infected in groups of size 10; 100; 1,000; 10,000; and even 100,000 (Weitz, 2020). The risk chart highlighted combinations of event size *n* and circulating cases in the United States that had equal risk. The visualized risk contours can be defined via a binomial statistical model as a set of values (*p,n*) such that risk is a constant *r* = 1 − (1-*p*)*^n^* (the implications of these risk contours at the early stages of the Covid-19 outbreak in the United States are discussed in (Arnold, 2020; Downey, 2020; Maggiacomo & Greshko, 2020)). Given a risk level *r* defined between 0 and 1, then the per-capita probability along an equirisk contour scales like 1/*n* (converging rapidly to 0 when n is large). Hence, large events can potentially seed transmission even when the per-capita probability that an individual is infected remains low.

With shelter-in-place orders now suspended in most of the United States, many businesses (from retail to sports), recreational facilities, daycare centers, schools (both K-12 and colleges/universities) are evaluating re-opening plans. These plans must also gauge likely risk. The *<u>Covid-19 Real-Time Event Risk Assessment</u>* website uses a data-driven approach to connect circulating case reports with risk assessment by adapting a binomial model of risk to real-time estimates at the county level. The central purpose is to quantify and visualize the expected risk associated with gatherings of different sizes and to help guide action-taking by policy makers, public health departments, as well as event planners and visitors. The interactive website has drawn over two million visitors since the launch of the county-level map on July 7, 2020; is updated daily, and continues to provide real-time estimates of event-associated risk. As we describe below, the visualized risk maps are intended to inform individuals on the need to take preventative steps to reduce new transmission, e.g., by avoiding large gatherings and wearing masks when in close contact with others.

## Results

### Real-time risk is heterogeneous, reflecting recent increases in cases

We utilized a binomial probability model to assess the risk that one or more individuals is infected with SARS-CoV-2 (see probability model; Methods). The risk that one or more individuals is infected at an event is heterogeneous. To assess variation, we measured county-level heterogeneity by combining time-varying estimates given reported case counts from May-August at the county level. Figure 1 shows four snapshots, spaced monthly, corresponding to estimated risk associated with a gathering of size 100 on the first of May, June, July, and August 2020. These snapshots reveals that gathering-associated risk was heterogeneous and concentrated in the Northeast (and to some extent in the Southwest) in early May with higher risk associated with the South and Southeast beginning in early June. Critically, the regional shift in current risk means that use of cumulative case or death counts do not necessarily provide near-term actionable information on ongoing risk. We note that estimates are affected by uncertainty in the ascertainment bias; the default option is 10x corresponding to the median of serologically positives to PCR positives in a locale-centered population surveys conducted in April-May 2020 (see Methods). In light of increased testing, we also include a 5x ascertainment bias (see Discussion for more details).

**Figure 1:**
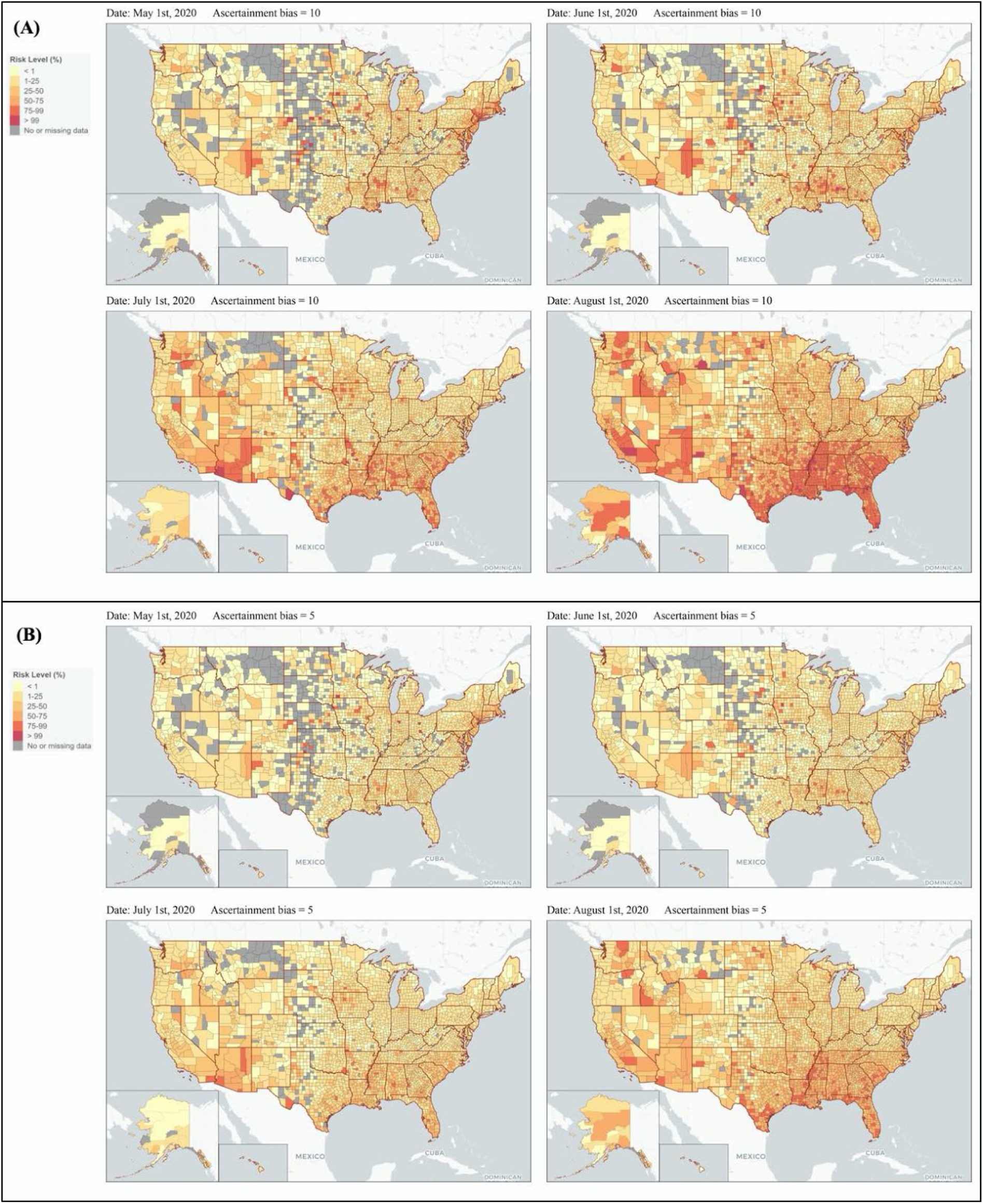
Heterogeneous risk map. The map depicts risk given events of size 50 using ascertainment biases of 10x (A) and 5x (B) on May 1, June 1, July 1 and August 1. Alaska and Hawaii were resized to be smaller than they actually are on the web.

### Information can be conveyed by focusing on risk associated with intermediate-scale events

One key choice in visually displaying risk is selecting event sizes that are meaningful in a public health context, can be pre-computed, and effectively communicate differential risk. Precomputation is key to accommodate a large number of users simultaneously. The choice of gathering size strongly influences the information content of county-level maps. The map includes six different colored bins representing the probability of an infected individual being present at an event: <1%, 1%-25%, 25%-50%, 50%-75%, 75%-99%, and >99%. We note estimation of risk as calculated via the binomial model which saturates at 1 when the size of the gathering *n* is much larger than *1/p*. For example, if *p* = 0.005 or 1 in 200, then events much larger than 200 will saturate near 1; in contrast, if *p* = 0.0001 or 1 in 10,000, then events much larger than 10,000 will saturate near 1. As a result, the map will be uniformly ‘light’ (associated with low risk) when events are sufficiently small and uniformly ‘dark’ (associated with high risk) when events are sufficiently large. This also suggests that displaying risk associated with intermediate size events will more effectively communicate differences between counties and states.

We used an information-based metric to assess the overall spatial heterogeneity of the county-level risk map. We denote the visual map information as the sum of −*q_i_*log(*q_i_*) where *q_i_* denotes the fraction of counties in the *i*-th risk category where *i* = 1 to 6 (per the number of data bins on the map). Note that small counties are weighted equally as large counties, and future work could use a population-weighted cartogram to allow users to visualize county areas in proportion to their respective populations. Figure 2 quantifies the information conveyed associated with visualizations across sizes from 10 to 1000 on August 1, 2020. The peak information is found at a size *n* = 70 and 142 for ascertainment biases 10x and 5x respectively, consistent with the maximum color divergence at intermediate risk sizes. This peak suggests that in early August, whereas most small events (of about 10) had relatively low risk everywhere and most large events (greater than 1000) had relatively high risk everywhere; the risk associated with intermediate sized events was strongly variable with region. Such variability is critical to informing opening decisions.

**Figure 2.**
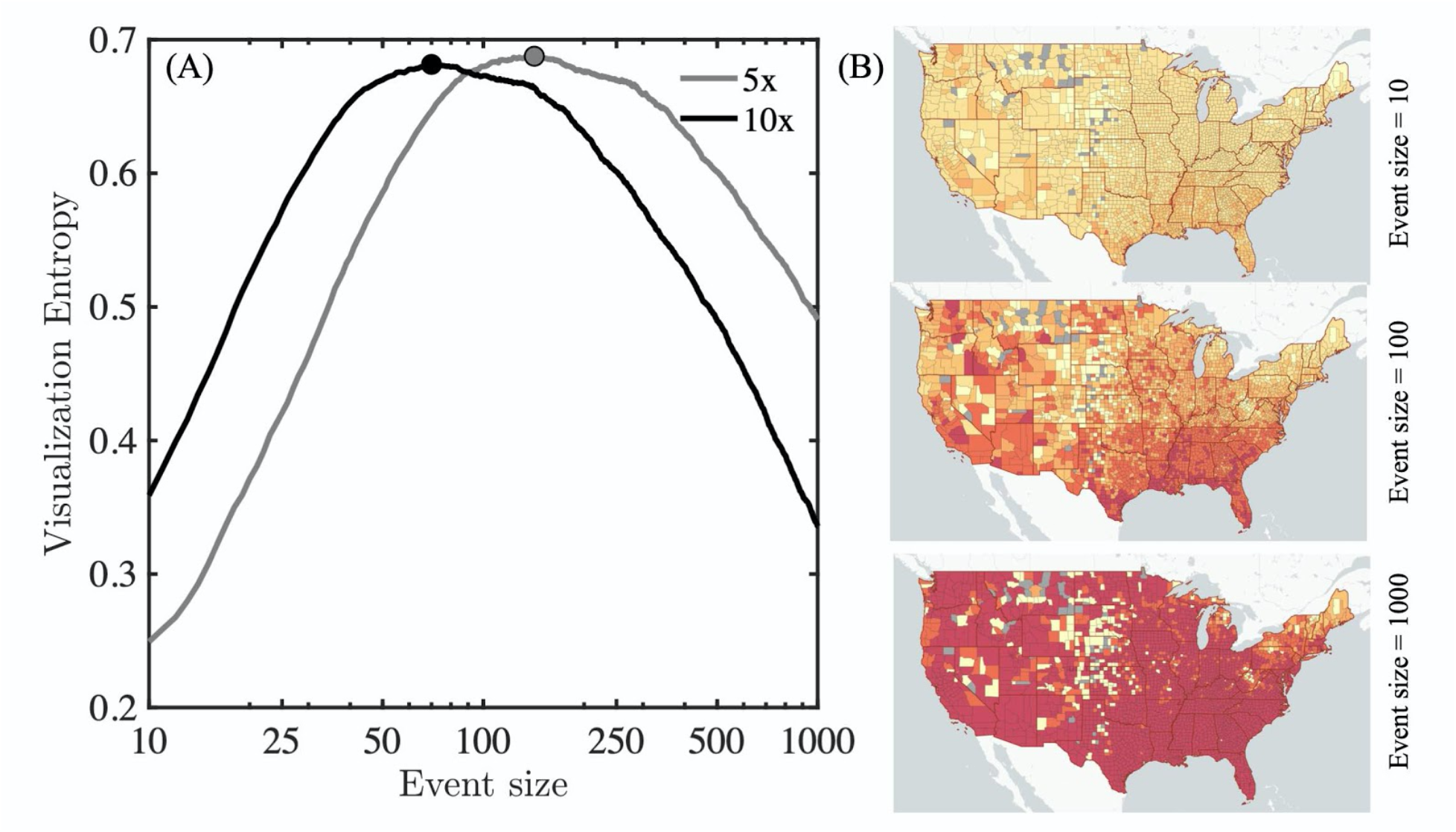
Visualizations of event-associated risk. An entropy-based index of heterogeneity in risk reveals that intermediate event sizes differentiate spatially heterogeneous risk as of August 1, 2020. (A) Visualization entropy as a function of event size using 5x and 10x ascertainment biases. (B) Maps illustrating that most counties appear to have similarly low risk when events are small (<10 individuals) or similarly high risk when events large (>1000 individuals). In contrast, the highest level of heterogeneity in risk is revealed given intermediate event sizes (50-150 individuals). Map visualizations use an assumption of 5x ascertainment bias.

### State-level variation in critical event sizes

The spatiotemporal variation in risk can be viewed a different way: by evaluating the location-dependent risk associated with a given event size. To do so, we fixed the event size as 50 and then estimated the state-level risk [1-(1-p)^50^]. Figure 3 arranges states as well as Washington DC and Puerto Rico in order of their relative risk effective August 15, 2020 from #1 (lowest state-level risk) to #52 (highest state-level risk). In many cases, states with high risk levels in May and June experienced declines throughout July and August, particularly in the Northeast. In contrast, states with lower risk levels in May and June experienced upsurges of cases (and risk) in July and August, especially in the South. This analysis further reinforces the spatiotemporal variation of event risk, as many states continue to have significantly elevated risk associated with gatherings of 50 (corresponding to a social gathering, bar, restaurant, business event or approximately two K-12 classes). This finding suggests that plans to reopen schools, colleges, and businesses should operate knowing that there is a significant risk of within-event transmission if precautions are not taken; and are robust to the choice of both ascertainment biases – either 5x or 10x.

**Figure 3.**
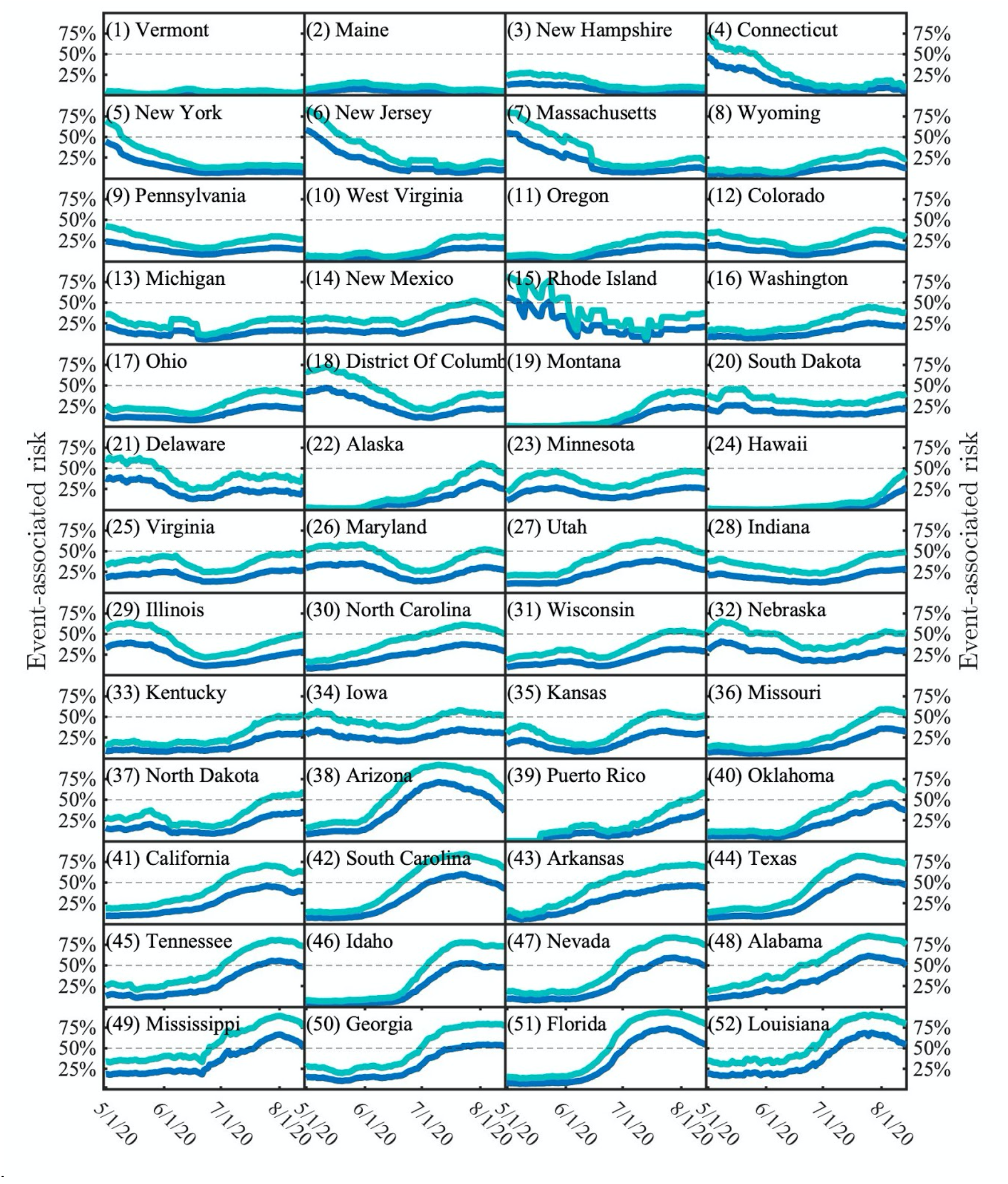
State-level risk associated with events of size 50 over time. The curves denote risk estimates assuming 5:1 (dark blue) and 10:1 (light blue) ascertainment biases. States are ordered as a function of ascending risk level as of August 14, 2020 (last point shown).

## Discussion

The *Covid-19 Event Risk Assessment Tool* provides real-time, localized information on risk associated with gatherings. The risk highlights the probability that one (or more) individuals may have Covid-19 in events of different sizes. By integrating real-time information aggregated via state health departments nationwide along with a simple statistical model, the website is able to capture, calculate, and disseminate information relevant to decision-making by the public that could help reduce risk and new transmission.

Static and interactive maps and as well as interactive data *dashboards*, i.e. sets of linked visualizations for data exploration (Yi et al., 2007), have proliferated since the start of Covid-19. Most dashboards allow visitors to choose one of four variables to display: number of cases, cases per capita (e.g. per 100,000 people), number of deaths and deaths per capita, for divisions within a single country, or for countries on a global map. Other behavioral maps have illustrated the reduction in mobility (GeoDS Lab @ UW-Madison, 2020) or polling results such as attitudes towards masks (Katz, Sanger-Katz, & Quealy, 2020). Like these maps, the *Covid-19 Event Risk Assessment Tool* describes relationship between disease spread and behavior; albeit in an effort to change rather than track behavior. This map is designed as a spatial decision support system (Armstrong & Densham, 1990) that allows the individuals to measure the risk of their own actions, and plan accordingly. It removes the burden of interpreting what case rates mean in a quantitative context by directly communicating a probability of encountering an infected individual via interactions. As a result, individuals can visualize themselves in a group, and decide whether this risk is worth taking.

The interpretable risk levels provided by the *Covid-19 Event Risk Assessment* website encourages visitors to take steps to reduce their risk of infection, such as physical distancing, washing hands, and wearing a mask. By illustrating how risk increases nonlinearly with event size, the tool may be particularly useful in encouraging large event planners to reschedule or cancel events, move to a safer format (e.g., outdoors where transmission risk is reduced or online when possible), averting potential exposures. As such, the website is of particular relevance given the relaxation of shelter in place orders across the United States, including restrictions on gatherings. These relaxation of non-pharmaceutical interventions imply that individuals must remain informed on the personal risk involved with everyday activities so as to modify their behavior accordingly.

There are multiple ways to extend these findings to improve local estimates. First, the model uses a binomial probability of risk that assumes that risk is homogeneous at county levels. We anticipate there will be variation within counties (e.g., see studies on heterogeneous risk within NYC boroughs (Sy et al., 2020)). However, because data on cases is reported at the county level, further refinement to tract or zip code levels is not yet feasible. In addition, the website does not break down risk in terms of other socioeconomic correlates, or by race, gender, or other personalized factors. Second, the risk model assumes that individuals are equally liked to attend an event; whereas increases in symptomatic case isolation implies that a fraction of infectious individuals are unlikely to attend events (the same applies to those hospitalized, albeit that is a much smaller fraction of the total). Yet, perhaps the largest driver of uncertainty remains ascertainment bias. Ascertainment bias denotes the number of actual cases for each documented case. A recent population-wide CDC serosurvey found that ascertainment bias ranged from 6-24 fold above PCR documented cases in March and April (Havers et al., 2020). Phase 2 serology surveys of populations revealed a range in ascertainment bias from between 2-14, with a median of 9x to 10x (Centers for Disease Control and Prevention, 2020a). Rapid, population-wide serosurveys are needed to connect case reports to localized estimates of ascertainment bias. Integration of such serosurveys at state levels or improvements in estimates of ascertainment bias (Perkins et al., 2020) could further refine event risk estimates on the website.

In closing, by connecting real-time case reports in the context of risk associated with events, the website attracted a significant visitor base, including more than two million visitors in the first two months after release of a county-wide risk tool. This interest showcases the importance of translating epidemiological statistics into real-world context. In doing so, we hope that health departments in the United States and globally consider integrating event-associated risk models in current and future pandemic responses as part of public awareness campaigns. Spatially resolved risk models can help to convey heterogeneous risk at local levels, and provide accessible information that can help to justify the choice of restrictions on gatherings as part of integrative campaigns to control spread. For SARS-CoV-2, the open-source and publicly available dashboard highlights the fact that there is a significant risk that one (or more) individuals may be infected in groups of 500-1000, irrespective of location as of mid-August 2020; these sizes are consistent with typical enrollment at K-12 schools. Hence, it is critical that re-openings of businesses and schools devise policies for testing, mask wearing, and other non-pharmaceutical interventions to ensure that one case does not soon become many.

## Methods

### Probability model

We estimated the probability that one or more individuals may have SARS-CoV-2 in events of different sizes via a binomial assumption of homogeneous risk. Let *p* denote the probability that a randomly selected individual in a focal population is infected. Hence, the probability that each of *n* individuals is not infected must be *(1-p)^n^* and by extension the probability that one (or more) individuals is infected must be *1-(1-p)^n^;* we define this as the event gathering risk. This formalism was used as the basis for early estimates to communicate risk of large gatherings in March 2020 using a scenario-based approach to estimating *p* within the United States (Arnold, 2020; Downey, 2020; Weitz, 2020).

### Circulating case estimate

At a county level, the circulating per-capita probability of infection is defined as the estimated number of circulating cases divided by the census population. The circulating case counts are defined, operationally, in two stages. First, the number of newly-documented cases over the past 10 days are obtained via data via state departments of public health. Data were aggregated and accessed from the New York Times’ repository of Covid-19 data (New York Times, 2020a) using a standard application programming interface. The choice of 10 days is consistent with CDC guidelines on durations of infectiousness (Centers for Disease Control and Prevention, 2020c). Second, the number of newly documented cases is multiplied by an ascertainment bias to yield the estimated number of circulating cases. The default ascertainment bias is 10x, consistent with a median of 9-10 in population-wide surveys conducted by the Centers for Disease Control and Prevention (Havers et al., 2020; Centers for Disease Control and Prevention., 2020a); with a secondary option of 5x.

### Visualization code

The code to visualize county- and state-level risk was written in R and used the R Shiny Package for map deployment. The input data was a county shapefile from the U.S. Census that included all 50 states, the District of Columbia, and Puerto Rico whose boundaries were generalized using the ‘rmapshaper’ package. This file was converted to a geojson file for faster drawing. The projection was relegated to a web Mercator standard instead of a traditional conic projection due to the constraints of the R package. New York City was agglomerated as a set of five counties in order to accommodate the New York Times’ county level case data (New York Times, 2020a), which reported New York City as a single region. The risk value shown on the county-level map takes into account the county’s new cases for the past 10 days, the user’s chosen ascertainment bias (5 or 10) from a radio button, and the user’s chosen case size from a slider with eight discrete increments (10, 25, 50, 100, 500, 1,000, 5,000, and 10,000). The map symbology was chosen as a univariate color ramp showing intensity in red, and allows for interactive zooming and panning. Upon hover, a pop up shows the county name and the likelihood (in terms of a percentage) that an individual at that event is infected with SARS-CoV-2.

### Web application

The web application is built using the R-Shiny web development framework and deployed as a self-contained Docker container using the open-source shiny-server. Application containers are deployed to a fleet of servers hosted at Georgia Institute of Technology, with multiple application instances running on each. Users are load-balanced across instances using Nginx. All static data used in the application (e.g. map HTML files, data used for interactive plots) are automatically updated and distributed to each application instance.

### Data Availability

Population demographics for US states and counties were obtained from the publicly available United States Census Bureau American Community Survey for 2018 (U.S. Census Bureau, 2019). State-level cases were obtained from CovidTracking.com - a project developed by Alexis Madrigal and colleagues at The Atlantic (The Atlantic Monthly Group, 2020). County-level cases were obtained from the New York Times github site (New York Times, 2020a).

### Code availability

The website code is open source and available on Github: https://github.com/appliedbinf/covid19-event-risk-planner.

## Data Availability

Data Availability: Population demographics for US states and counties were obtained from the publicly available United States Census Bureau American Community Survey for 2018 (U.S. Census Bureau, 2019). State-level cases were obtained from CovidTracking.com, a project developed by Alexis Madrigal and colleagues at The Atlantic (The Atlantic Monthly Group, 2020). County-level cases were obtained from the New York Times github site (New York Times, 2020a). 
Code availability: The website code is open source and available on Github: https://github.com/appliedbinf/covid19-event-risk-planner. 

https://github.com/appliedbinf/covid19-event-risk-planner

## Acknowledgements

Research effort by JSW was enabled by support from grants from the Simons Foundation (SCOPE Award ID 329108), the Army Research Office (W911NF1910384), National Institutes of Health (1R01AI46592-01), National Science Foundation (1806606, 1829636, and 2032084), and additional support from the Charities Aid Foundation and The Marier Cunningham Foundation. Research effort by CA was enabled by the College of Design and College of Computing at the Georgia Institute of Technology.

## Author contributions

A.C. led the development of the web server, contributed analysis, and analyzed data. S.L. led the development of the map-based visualizations, contributed analysis, and analyzed data. M.H. contributed to website design, study design, and edited the manuscript. T.H. contributed to the development of the web server. C.A. co-designed the study, provided oversight to all aspects of map development, analyzed data, and co-wrote the manuscript. J.S.W. designed the study, provided oversight for all aspects of the study, developed the core modeling framework, analyzed data, and wrote the manuscript.

